# The impact of the number and the size of clusters on prediction performance of the stratified and the conditional shared gamma frailty Cox proportional hazards models

**DOI:** 10.1101/2025.10.17.25338219

**Authors:** Daniele Giardiello, Edoardo Ratti, Peter C. Austin

## Abstract

Researchers in biomedical research often analyse data that are subject to clustering. Development and validation of risk prediction models generally assumes independence of observations. For survival outcomes, the Cox proportional hazards regression model is commonly used to estimate an individual’s risk at fixed time horizons. The stratified Cox proportional hazards and the shared gamma frailty Cox proportional hazards regression models are two common approaches to account for the presence of clustering in the data. The accuracy of the predictions of these two approaches has not been examined. We conducted a set of Monte Carlo simulations to assess the impact of the number of clusters, the size of the clusters, and the within-cluster correlation in outcomes on the accuracy of the conditional predictions developed using the stratified and the shared gamma frailty Cox proportional hazards regression model. We compared the accuracy of the predictions in terms of discrimination, calibration and overall performance metrics. We found that the stratified and the shared gamma frailty model had similar performance, especially for larger size and higher number of clusters. For small cluster size, we observed slightly better discrimination and overall performance for the stratified model and better calibration for the shared gamma frailty model at shorter prediction horizons. The utility of the stratified Cox proportional hazards model for risk prediction is limited especially for high within-cluster correlation and when clusters are small, and at longer prediction horizons. Our results were accompanied with two applications using open source data on myelodysplastic syndrome and bladder cancer.

## 1. Introduction

Researchers in biomedical research frequently analyse data that are subject to clustering. Over the last decades, international collaborations have become more frequent, allowing researchers to investigate disease diagnosis and prognosis in patients from different centres and countries. Multicentre clinical trial data, observational data with geographic clusters, family-based studies, and individual patient data meta-analysis are examples of individuals nested within different clusters such as centres, countries, genetically related individuals and studies, respectively.

An important aim of clinical risk prediction models is to predict an individual’s risk of the outcome using statistical regression models that typically assume independence among observations. Such ordinary assumption implicitly consider that populations are homogeneous after taking into account all relevant covariates: all individuals with the same individual characteristics included in the risk prediction model have the same estimated predicted risk. Despite tight study protocols, variation in hospital- or country-specific clinical practice may occur, leading to centre-to-centre heterogeneity in outcomes. This heterogeneity cannot be explained by the information included in the model due to financial costs or because all relevant information is not (yet) known. Centre-, or country-specific heterogeneity is common in medical studies. Good clinical examples are represented by haematological cancers where hematopoietic cell transplantation requires adequate infrastructure, personnel and resources at transplant centres^1^. The “centre effect” has been commonly investigated in the haematological malignancies literature in the last decades^2–4^. Thus, when observations are nested within clusters, regression models should consider the presence of clustering in the data used to develop, to validate, and to deploy risk prediction models

Logistic regression and the Cox proportional hazards model are frequently used to develop prediction models for the analysis of binary and time-to-event outcomes in biomedical and epidemiological research^5,6^. For prediction models using logistic regression, the impact of clustering has been more largely investigated^7,8^. Less is known about prediction models for time-to-event outcomes using clustered data. The Cox proportional hazards model allows one to estimate an individual’s risk of experiencing the outcome at specific time horizons such as, for example, to estimate the 10-year risk of death. The stratified Cox proportional hazards models and frailty Cox models are the most common survival models that account for clustering^9^. Both approaches treat potential correlation of survival times of individuals within a cluster in different way. The stratified Cox proportional hazards models assume that individuals in each cluster (namely, strata) have different baseline hazard functions. Using a different baseline hazard function for each cluster, the estimated risk predictions can be different in individuals with the same characteristics coming from different clusters. Frailty Cox proportional hazards models include random effects assuming that the clusters such as centres or countries are drawn from a population of possible clusters. This enables one to account for unobserved heterogeneity between clusters. For computational and mathematical reasons, the most common frailty Cox proportional hazards model is the shared gamma frailty distribution^9,10^. Risk predictions can be obtained in closed mathematical form using the shared gamma frailty Cox proportional hazards model. Both conditional and marginal predictions consider clustering in predicting individual’s risk. In particular, the conditional frailty models estimate an individual’s risk among patients from the same clusters that were used to develop the model. The marginal frailty models estimate predictions that are averaged across clusters. Thus, predictions can also be also estimated for patients coming from different clusters than those used to develop the model considering that data are nested within clusters.

To the best of our knowledge, there is a paucity of research on the impact of the size and the number of clusters on the accuracy of predictions obtained from the stratified and the conditional frailty Cox proportional hazards models. The objective of the current paper was to examine the impact of the number of clusters, the cluster size, and the within-cluster correlation in outcomes on the accuracy of the conditional prediction models developed using the stratified and frailty Cox proportional hazards regression model. The paper is structured as follows: in Section 2, we describe a series of Monte Carlo simulations that were designed to address this question. In Section 3, we report the results of these simulations. In Section 4 and 5, we provide two case studies to illustrate the application of these methods. In the first case study we use data from the Center for International Blood & Marrow Transplant Research (CIMBTR), in the second case study we use bladder cancer data extracted from the European Organization for Research and Treatment of Cancer trials. Finally, in Section 6, we summarize our findings and place them in the context of the existing literature.

## 2. Monte Carlo simulations methods

We conducted a set of Monte Carlo simulations to examine the impact of the size of clusters, the number of clusters, and within-cluster correlation on the accuracy of predictions obtained from the stratified and the shared gamma frailty Cox proportional hazards models using clusters as stratification and frailty term, respectively. We simulated clustered data with time-to-event outcomes related to subject-level covariates.

### 2.1 Factors in the Monte Carlo simulations

We allowed three factors to vary in the Monte Carlo simulations: (i) the number of clusters (N_cluster_); (ii) the mean number of subjects per cluster (N_subjects_); (iii) the within-cluster correlation in the time-to-event outcomes (*τ*). The first took on values from 2 to 10 in increments of 2, for a total of 5 values. The second took on four values: 25, 50, 100, 200, for a total of 4 values. The third took on four values: 0.01, 0.05, 0.10, and 0.20, for a total of 4 values. We used a full factorial design and thus examined 80 different scenarios. In each scenario we generated 1 000 datasets.

### 2.2 Data Generating Process

For each of the N_cluster_ clusters we generated a random variable denoting the size of the cluster (N_subjects_). The size of each cluster was drawn from a uniform distribution with the lower and upper limit of 0.5*N_subjects_ and 1.5*N_subjects_, respectively. Thus, the average size of each cluster was N_subjects_, rounded to the nearest integer. In every scenario, we generated two independent subject-level baseline covariates: X1 and X2. The first was continuous and was generated from a standard normal distribution, while the second was binary and was generated from a Bernoulli distribution with parameter 0.5. Thus, in expectation, Pr(X2 = 1) = 0.5.

For each subject we simulated a time-to-event outcome and a censoring time. First, we simulated correlated individual-level time-to-event outcomes using a gamma frailty model combined with Bender and colleagues’ approach for simulating time-to-event outcomes using a Cox–Weibull model^11^. Define *LP_ij_* to be the log-hazard of the outcome, which is a linear combination of the two covariates (i.e., X1 and X2) and a cluster-specific random effect *µ*:

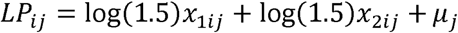

where the subscript *ij* denotes the *i^th^* subject in the *j^th^* cluster. The cluster-specific random effects *z_j_* = *exp*(*µ_j_*) are randomly drawn from a gamma distribution with shape parameter 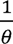 and scale parameter *θ* where 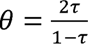. The rationale for this choice of shape and scale parameter is that Kendall’s *τ* is used as a measure of dependence for the gamma frailty model assuming values between zero (no within-cluster correlations) and one (the highest within-cluster correlation)^9,10,12^. For a specified value of *θ*, we have 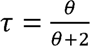. Thus, we can induce data with a specific degree of within-cluster dependence for a specified value of *θ*. We then generated time-to-event outcomes under the Cox-Weibull model: *T_ij_* = 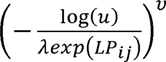 where *u* is drawn from a standard uniform distribution, and *λ* = 0.01 and *v* = 0.75. For each subject we also simulated independent censoring times from an exponential distribution. The observed survival time was defined to be the minimum of the event time and the censoring time. The rate parameter of the exponential distribution was chosen using an iterative bisection procedure so that 20% of the simulated subjects would be censored^13^. We repeated this entire process with the same cluster size and frailty terms to generate validation samples (which is equivalent to bisect a sample of 2*N_subjects_ per cluster for every scenario). Thus, we evaluated the predictive performances of the stratified and the frailty Cox proportional hazards models fitted using the derivation samples and evaluated in the validation samples (see the subsequent section).

After we had the simulated an event time for every scenario for each individual, we generated a super-population with 1 000 clusters and 1 000 subjects per cluster for a total of 1 000 000 subjects for each *τ* for a total of 4 000 000 subjects. We determined the *25^th^, 50^th^, 75^th^*percentiles of the event times. These will be used as the prediction horizons at which predicted event probabilities will be estimated from the stratified and the frailty Cox proportional hazards models (see the next section). We refer to these three quantities of event time as *T_25_*, *T_50_* and *T_75_*.

For each of the 80 different scenarios, we generated 2 000 random samples (1 000 for derivation sample and 1 000 for validation sample), for a total of 160 000 random samples.

### 2.3 Statistical analyses

In each of the 80 000 random derivation samples (1 000 derivation samples for 80 different scenarios), we fit a stratified and a shared gamma frailty Cox proportional hazard model in which the hazard of the outcome was regressed on the two subject-level covariates including clusters as a stratified or as a frailty term, respectively. The fitted models were then applied to the validation samples and a predicted probability of the outcome at *T_25_*, *T_50_* and *T_75_* was obtained for each individual in the corresponding validation sample.

We used the following quantitative metrics to evaluate the prediction performances of the models: for discrimination performances, the time-varying Area Under the receiver operating characteristic (ROC) Curve, which we refer to as time-varying AUC; for calibration performances, the integrated calibration index (ICI), E50, E90, and the ratio of observed-to-predicted risk^14–16^. We also computed the Index of Prediction Accuracy (IPA), which is equivalent to the scaled Brier score, at each of the prediction horizons (strictly speaking the IPA is not a measure of only discrimination, but combines discrimination and calibration; we include it here for the sake of completeness)^17^. The ICI, E50, and E90 are the mean, median, and *90^th^* percentile, respectively, of the absolute difference between predicted survival probabilities and smoothed survival frequencies^18^. Smoothed survival frequencies were obtained using Kooperberg’s flexible adaptive hazard regression model with the complementary log–log transformation of the predicted probabilities as the sole predictor variable^19^. The ratio of observed-to-expected risk was computed by dividing the observed risk of the outcome at the specified time horizons by the mean predicted risk of the outcome derived from the fitted Cox regression models. The observed risk of the outcome was determined using the Kaplan–Meier estimate of risk at the given duration of time. Thus, in each simulated validation sample, we computed the time-varying AUC, time-varying IPA, ICI, E50, E90, the ratio of observed-to-expected risk. The mean and standard deviation of each of these measures were then computed across the 1 000 simulated validation samples for each simulation scenario. These measures represented the Monte Carlo estimates.

### 2.4 Software

The simulations were conducted using the R statistical programming language (version 4.3.3)^20^. The Cox model was fit using the coxph function from the survival package (version 3.8-3). The penalized partial likelihood estimation is used to estimate coefficients for the gamma frailty model through the coxph function from the survival package including the frailty term^21,22^. Observed risk for the observed-to-expected ratio was computed using the survfit function from the survival package. The predicted risk from the fitted stratified Cox model was estimated using the predictSurvProb function from the pec package (version 2025.06.24). The time-varying AUC and IPA were computed using the Score function from the riskRegression package (version 2025.05.20). The ICI, E50, and E90 were computed using the smoothed survival frequencies generated through the hare function from the polspline package (version 1.1.25). Simulation results were summarized using the simsum function from the rsimsum package (version 0.13.0).

## 3. Monte Carlo simulations results

We report our results separately for different time horizons *T_25_*, *T_50_* and *T_75_* (Q1/Q2/Q3 for the three quantiles of event times in the super-population).

The simulation results are summarized in Figure 1 for *T_25_,* Figure 2 for *T_50_,* Figure 3 for *T_75_*. Each figure consists of six panels, one for each of the performance metrics. Each panel uses a nested loop plot to report the performance metric across the different scenarios^23^. The value of the performance metric is reported on the vertical axis. At the top of each panel are three black step functions, one for each of the factors: the average number of subjects per cluster (i.e., the size of the cluster, N_subjects_), the number of clusters (N_clusters_), and the homogeneity within clusters *τ* (tau). Below these three black step functions are two step functions (in red for the stratified Cox proportional hazard model and in blue for frailty Cox proportional hazard model). For a given combination of size, number of clusters and homogeneity within clusters, one determines the vertical line that bisects each of the top three black step functions at the given combination of values. The vertical location where this vertical line bisects the step functions for the stratified and the frailty Cox model is the value of the performance metric for that model and for that scenario. In interpreting the results, recall that higher values of AUC and IPA denote improved model performance, while lower values of ICI, E50, and E90 denote improved model performance. For the observed-to-expected ratio, a better performing model will have a ratio closer to one.

**Figure 1.**
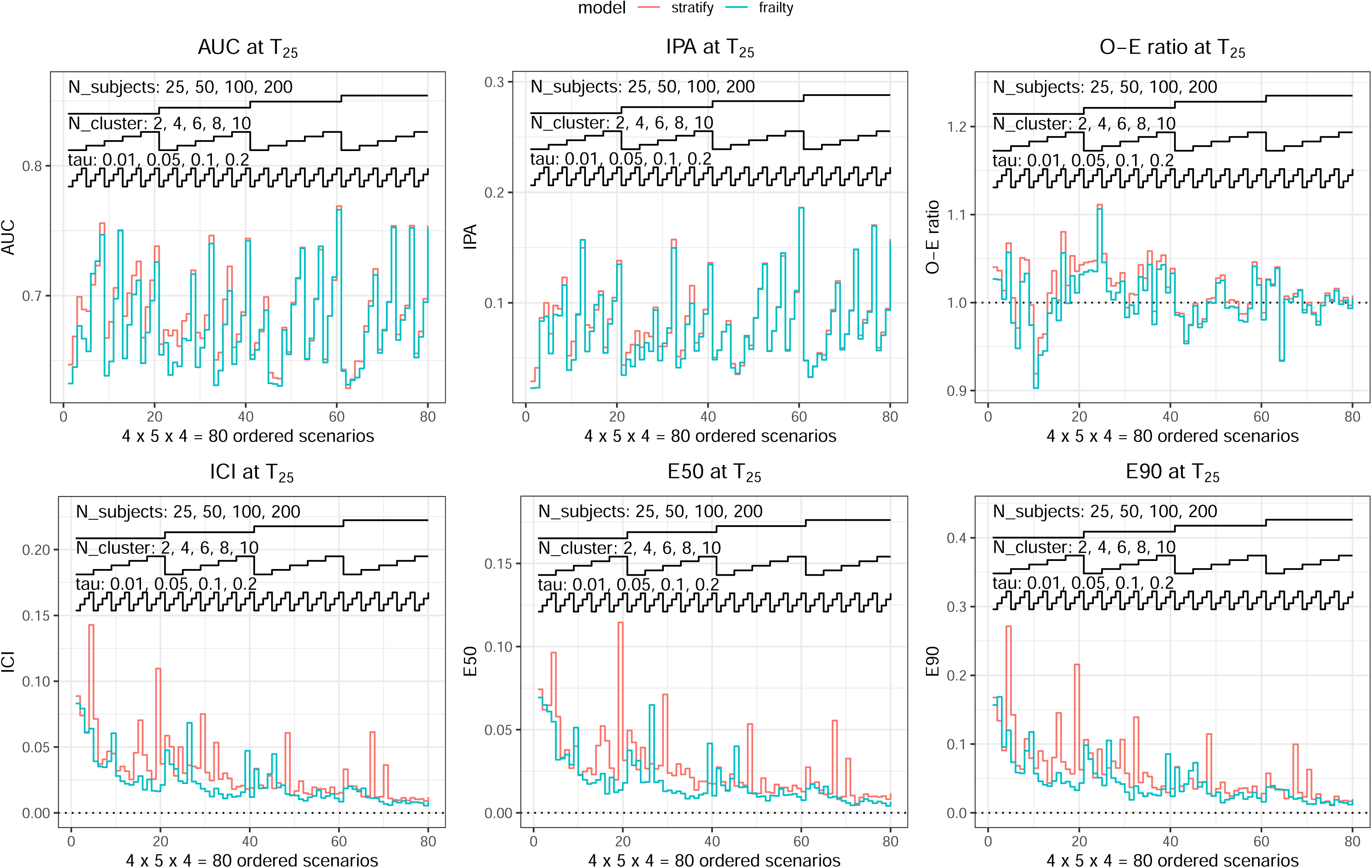
Prediction performances at T_25_.

**Figure 2.**
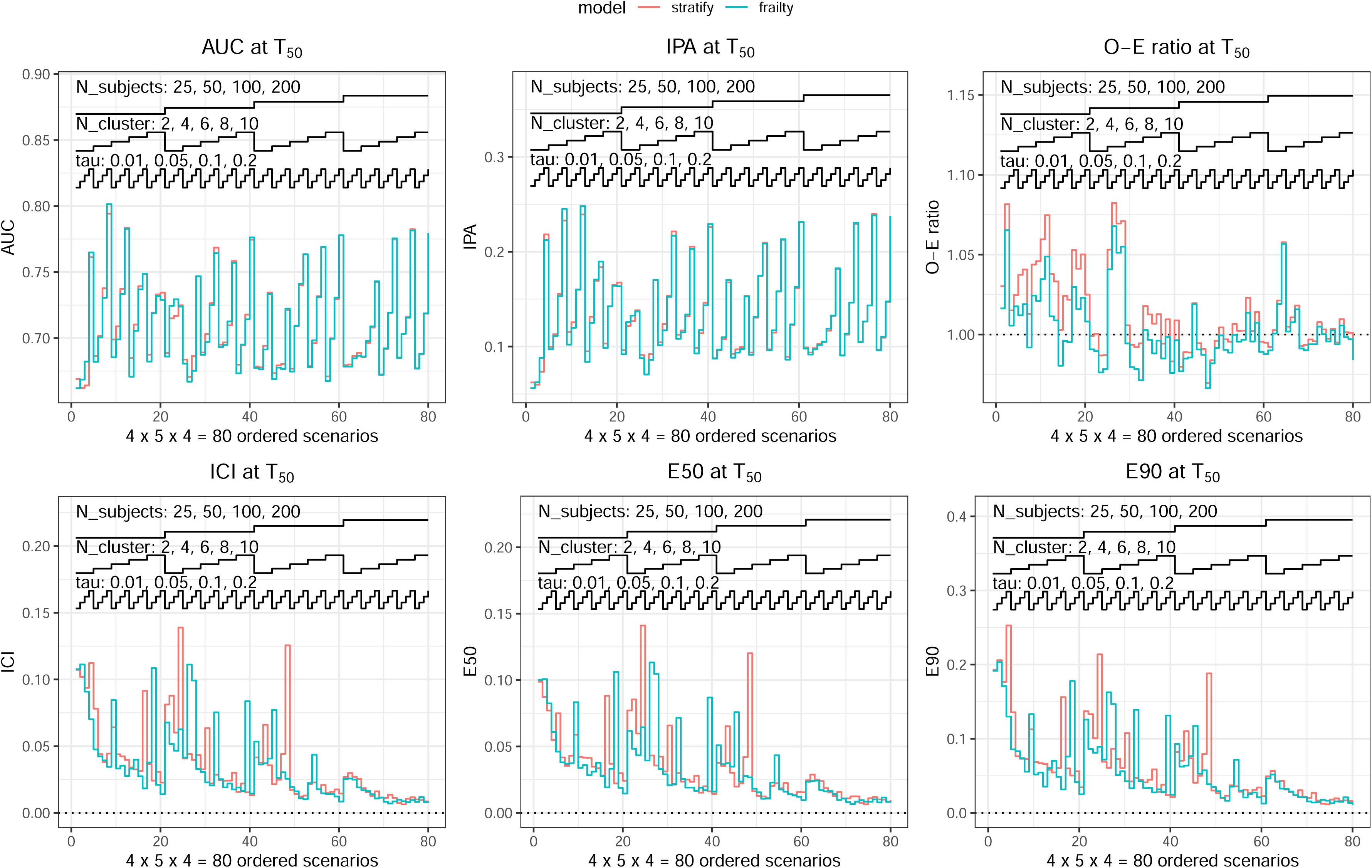
Prediction performances at T_50_.

**Figure 3.**
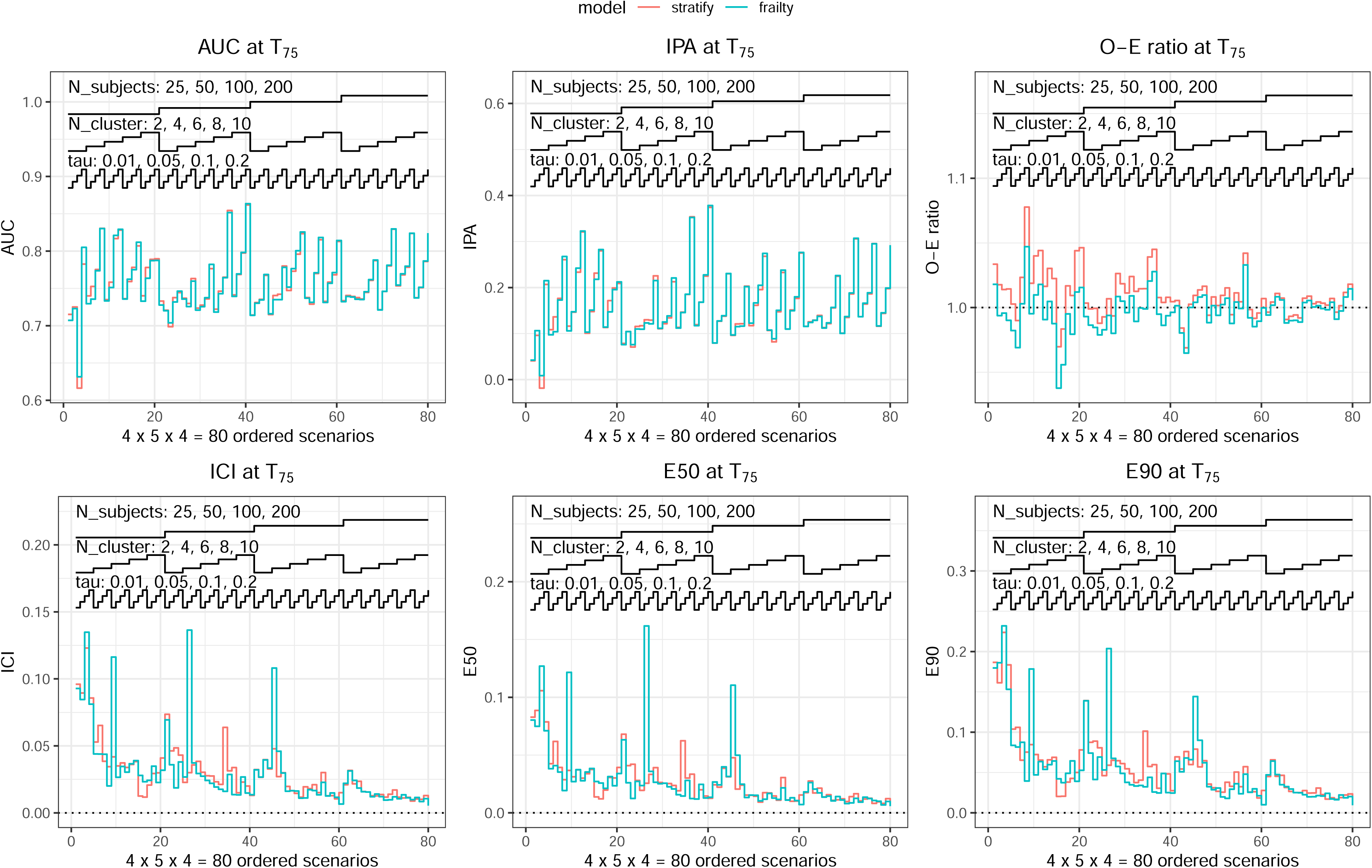
Prediction performances at T_75_.

The use of the stratified Cox model resulted in predictions at *T_25_* with slightly higher AUC compared to the frailty model in settings with smaller cluster size and especially low homogeneity within clusters (Figure 1 top left panel). The difference in AUC was attenuated for larger cluster size. Differences between the two models tended to be negligible with higher homogeneity within clusters and longer time horizons (Figure 1-2-3 top left panels). However, the stratum-specific (i.e., cluster-specific) baseline hazard from the stratified Cox models could not be estimated for clusters in which the time horizons were greater than the maximum event time in the derivation sample. Thus, it was impossible to estimate the corresponding predictions from the stratified Cox models. This limited the comparisons between the two models: predictions from the stratified Cox model could not be estimated especially in scenarios with longer time horizons, higher within-cluster homogeneity, and particularly for small cluster sizes and higher number of clusters. Among the evaluable comparisons, the AUC difference between the stratified versus frailty models across all scenarios ranged from −0.002 to 0.017 with a median of 0.002 (25^th^ and 75^th^ percentiles: 0.001 and 0.006) (Table 1).

**Table 1:** comparison of stratified versus frailty Cox proportional hazards models on prediction performance metrics across all comparable simulation scenarios and across all time horizons.

| **Performance metric** | **Comparison** | **Minimum** | **25<sup>th</sup> percentile** | **50<sup>th</sup> percentile** | **75<sup>th</sup> percentile** | **Maximum** |
| --- | --- | --- | --- | --- | --- | --- |
| AUC | Stratify vs frailty | -0.002 | 0.001 | 0.002 | 0.006 | 0.017 |
| IPA | Stratify vs frailty | -0.011 | 0.000 | 0.001 | 0.004 | 0.013 |
| O-E ratio | Stratify vs frailty | -0.019 | -0.002 | 0.003 | 0.012 | 0.028 |
| ICI | Stratify vs frailty | -0.069 | 0.000 | 0.002 | 0.009 | 0.050 |
| E50 | Stratify vs frailty | -0.081 | 0.000 | -0.001 | 0.003 | 0.046 |
| E90 | Stratify vs frailty | -0.096 | -0.001 | 0.005 | 0.017 | 0.106 |

AUC: Area Under the receiver operating characteristic Curve; O-E: observed-to-expected; ICI: Integrated Calibration Index

Both models resulted in estimates with similar IPA (Figure 1-2-3 top centre panels). The stratified Cox model resulted in slightly higher IPA at *T_25_* especially for small cluster size (Figure 1 top centre panel). For every scenario, the number of comparisons between the two models decreased with longer time horizons and higher homogeneity within clusters (especially for higher number of clusters with smaller size) for the reason described above. Among the evaluable comparisons, the IPA difference between the stratified versus frailty models across all scenarios ranged from −0.011 to 0.013 with a median of 0.001 (25^th^ and 75^th^ percentiles: 0.000 and 0.004) (Table 1).

The frailty Cox model tended to result in the observed-to-expected ratio slightly closer to one compared with the stratified Cox model when clusters were simultaneously characterized by small size, low number of clusters, and low homogeneity especially at *T_25_* (Figure 1-2-3 top right panels). The frailty Cox model tended to results in the observed-to-expected ratio closer to one compared to the stratified Cox model for most of the scenarios with small cluster size at longer time horizons (Figure 2-3 top right panels). The difference in observed-to-expected ratio was attenuated for larger cluster size. For every scenario the number of comparisons between the two models decreased with longer time horizons and higher homogeneity within clusters (especially for higher number of clusters with smaller size). Among the evaluable comparisons, for each observed-to-expected ratio we computed the absolute value of its difference from 1 and then computed the difference between these absolute differences. The observed-to-expected differences between the stratified versus frailty models across all scenarios ranged from −0.019 to 0.028 with a median of −0.003 (25^th^ and 75^th^ percentiles: −0.002 and 0.012) (Table 1).

The use of the frailty Cox model tended to result in predictions that provided better calibration measures (ICI, E50, E590) compared to the stratified Cox in several scenarios with either smaller cluster size, low homogeneity within clusters, or short time horizon (Figure 1-2-3 bottom panels). The difference of the two models in terms of calibration attenuated as size and number of clusters increased especially at longer time horizons (Figure 1-2-3 bottom panels). However, for every scenario, the number of comparisons between the two models decreased with longer time horizons and higher homogeneity within clusters (especially for higher number of clusters with smaller size). Among the evaluable comparisons, the ICI difference between the stratified versus frailty models across all scenarios ranged from −0.069 to 0.050 with a median of 0.002, (25^th^ and 75^th^ percentile: 0.000 and 0.009). The corresponding E50 and E90 differences between the two models ranged between −0.081 and 0.046 and between −0.096 and 0.106, respectively (Table 1). The frailty Cox model tented to be more efficient than the stratified Cox at short time horizons especially for calibration measures (see supplementary). Details about comparisons between the prediction performances of the two models across all scenarios by different time horizons are provided in the supplementary.

## 4. Application to mortality after transplant in myelodysplastic syndrome patients using the CIMBTR data

We provide an empirical analysis comparing the performance of the stratified and the shared gamma frailty Cox proportional hazards models to predict overall survival in patients with myelodysplastic syndrome (MDS) undergoing allogenic hematopoietic cell transplantation (HCT) using the CIMBTR data. The CIMBTR is a combined research program for the Medical College of Wisconsin and the National Marrow Donor Program. The CIBMTR comprises a voluntary network of more than 450 transplantation centres worldwide that contribute data on consecutive allo and autologous HCTs to a centralized statistical centre. Observational studies conducted by the CIBMTR are performed in compliance with all applicable federal regulations pertaining to the protection of human research participants. Protected health information used in the performance of such research is collected and maintained in the capacity of the CIBMTR as a public health authority under the Health Insurance Portability and Accountability Act Privacy Rule. Additional details are provided elsewhere^24^.

### 4.1 Methods

We used data with patients diagnosed with MDS enrolled in the CIMBR research database from 2004 to 2015. All patients underwent human leukocyte antigen (HLA) – matched mismatched-donor or umbilical cord transplant. Details are described by Nazha et al. ^25^. The dataset, consisted of 1,503 patients from 125 centres, was used to develop a prediction model for time-to-event outcomes after HCT such as overall and relapse-free survival and to compare the new proposed model with other international recognized prognostic scoring systems: the International Prognostic Scoring System (IPSS), its revised version (IPSS-R), and the CIMBR-MDS scores^26–28^. All scoring systems combined different patients and donors’ clinical, haematological and genetics characteristics without considering data clustered in different transplantation centres.

We used the Monte Carlo cross-validation procedure to assess prediction performances of the stratified and frailty Cox model using the transplantation centres as clusters. One hundred repeated random splits were used to define the derivation and validation samples. We stratified the repeated random splits by the transplantation centre (i.e., the cluster) to obtain the same centres in both the derivation and validation samples. Thus, in each centre, we split the observations in the derivation and validation samples to preserve the homogeneity within cluster on both derivation and validation samples. The splitting proportions were fixed at 70% / 30% to generate the derivation and validation samples in every cluster, respectively. We excluded centres with less than 10 patients since it was not possible to obtain sufficient patients in both derivation and validation sample. We developed the prediction models using the stratified and the shared gamma frailty Cox models across all 100 random derivation samples with the CIMBR-MDS prognostic score as predictor to estimate the overall mortality at 12, 24 and 36 months. Individuals missing of the CIMBR-MDS prognostic score were excluded. A total of 631 patients (369 events) from 28 centres were included. The numbers of patients per centre varied from 10 to 65, with mean 22.5 and median 16.5. We assessed the prediction performances in the 100 random validation samples at the three time horizons (12, 24, and 36 months) using the following metrics: time-varying AUC, time-varying IPA, observed-to-expected ratio, ICI, E50 and E90. The mean of the performances metrics across the 100 random validation samples with the corresponding standard deviation were reported at each prediction time horizon to assess the prediction performances of the stratified and the frailty Cox models. The accompanying R code for the case study is available in the supplementary online material and at https://github.com/danielegiardiello/ClusterSurvPred

### 4.2 Results

Results are reported in Table 2. The results suggest very similar prediction performances between the stratified and the frailty models. Discrimination performance measures were on average slightly better for the stratified model. IPA and calibration performances were slightly better for the frailty model. Not all the repeated random splits were usable for the stratified models due to the impossibility to estimate the baseline hazard for clusters (and the corresponding predictions) in which the time horizons were greater than the maximum event time observed in the derivation sample: 99, 95 and 37 out of 100 random splits were useable at 12, 24 and 36 months, respectively.

**Table 2:** accuracy of the predictions for the case study by model and time horizon for mortality after transplant in myelodysplastic syndrome patients using the CIMBTR data example.

| Time horizon | Model | AUC<br>(sd) | IPA<br>(sd) | O-E ratio<br>(sd) | ICI<br>(sd) | E50<br>(sd) | E90<br>(sd) | % usable<br>random<br>splits |
| --- | --- | --- | --- | --- | --- | --- | --- | --- |
| 12 months | Stratified | 0.623<br>(0.037) | 0.009<br>(0.049) | 1.040<br>(0.103) | 0.092<br>(0.050) | 0.077<br>(0.046) | 0.183<br>(0.046) | 100% |
|  | Frailty | 0.619<br>(0.031) | 0.045<br>(0.032) | 0.994<br>(0.094) | 0.059<br>(0.058) | 0.050<br>(0.059) | 0.112<br>(0.059) | 99% |
| 24 months | Stratified | 0.641<br>(0.031) | 0.024<br>(0.052) | 1.040<br>(0.087) | 0.087<br>(0.038) | 0.077<br>(0.046) | 0.167<br>(0.033) | 95% |
|  | Frailty | 0.608<br>(0.039) | 0.031<br>(0.039) | 0.995<br>(0.088) | 0.065<br>(0.056) | 0.050<br>(0.059) | 0.124<br>(0.057) | 100% |
| 36 months | Stratified | 0.664<br>(0.035) | 0.054<br>(0.051) | 1.040<br>(0.072) | 0.087<br>(0.038) | 0.079<br>(0.081) | 0.158<br>(0.081) | 37% |
|  | Frailty | 0.631<br>(0.040) | 0.053<br>(0.042) | 1.000<br>(0.080) | 0.065<br>(0.035) | 0.057<br>(0.034) | 0.127<br>(0.034) | 100% |

sd: standard deviation; AUC: Area Under the receiver operating characteristic Curve; O-E: observed-to-expected; ICI: Integrated Calibration Index

## 5. Application to bladder cancer data

We also applied the same analysis framework described previously using data on bladder cancer patients participating in a European Organization for Research and Treatment of Cancer (EORTC) trial ^29^. We used the free available data bladder from the R package frailtyHL consisting of 396 patients from 21 cancer centres. As described in the previous paragraph, we excluded centres with less than 10 patients to assess the prediction performances of the stratified and the shared frailty gamma Cox models to estimate disease-free survival at 12, 24 and 36 months using the Monte Carlo cross-validation procedure. The following predictors were included: age, sex, chemotherapy, number of tumours, tumour size, T stage and grade. A total of 355 patients from 14 centres were included. Two hundred and fifty-three events were observed (175 recurrences and 78 deaths before recurrence). The number of patients per centre varied between 11 and 78 with mean 27.3 and median 18. The accompanying R code for the case study is available in the online supplementary material and at https://github.com/danielegiardiello/ClusterSurvPred.

### 5.1 Results

Results are reported in Table 3. The results suggest similar prediction performances. All performance measures were slightly better for the frailty model. Not all the repeated random splits were usable for the stratified models: 99 out of 100 sample were usable at 12 and 24 months, while 91 at 36 months.

**Table 3:** accuracy of the predictions for the case study by model and time horizon for the bladder cancer data example.

| Time horizon | Model | AUC<br>(sd) | IPA<br>(sd) | O-E ratio<br>(sd) | ICI<br>(sd) | E50<br>(sd) | E90<br>(sd) | % usable<br>random<br>splits |
| --- | --- | --- | --- | --- | --- | --- | --- | --- |
| 12 months | Stratified | 0.623<br>(0.050) | 0.019<br>(0.062) | 1.040<br>(0.148) | 0.126<br>(0.078) | 0.113<br>(0.078) | 0.239<br>(0.128) | 100% |
|  | Frailty | 0.634<br>(0.049) | 0.044<br>(0.043) | 0.985<br>(0.107) | 0.101<br>(0.066) | 0.090<br>(0.068) | 0.191<br>(0.104) | 99% |
| 24 months | Stratified | 0.605<br>(0.046) | -0.015<br>(0.070) | 1.050<br>(0.129) | 0.131<br>(0.069) | 0.120<br>(0.071) | 0.246<br>(0.114) | 95% |
|  | Frailty | 0.616<br>(0.046) | 0.024<br>(0.050) | 1.010<br>(0.122) | 0.103<br>(0.065) | 0.093<br>(0.067) | 0.189<br>(0.101) | 100% |
| 36 months | Stratified | 0.565<br>(0.049) | -0.058<br>(0.075) | 1.040<br>(0.109) | 0.141<br>(0.070) | 0.131<br>(0.069) | 0.265<br>(0.116) | 91% |
|  | Frailty | 0.594<br>(0.049) | 0.001<br>(0.054) | 0.996<br>(0.128) | 0.104<br>(0.068) | 0.093<br>(0.070) | 0.192<br>(0.104) | 100% |

sd: standard deviation; AUC: Area Under the receiver operating characteristic Curve; O-E: observed-to-expected; ICI: Integrated Calibration Index

## 6. Discussion

Large electronic health records from different data sources have become more common in the last decades. The use of individual data from multiple sources, also better known in medical research as individual patient meta-analysis (IPD-MA), are more commonly encouraged and used to develop and evaluate risk prediction models considering data from different studies^30–32^. In this work we used simulations to investigate the impact of the size, number of clusters and within-cluster homogeneity on risk prediction performances of the Cox proportional hazards models for observations nested within a cluster such as a study centre. We summarize our findings as follows: first, the stratified Cox model showed slightly better discrimination compared to the frailty Cox model where the number, the size of the clusters and homogeneity within clusters were small. AUC were very similar as the number and size of clusters increased. Second, IPA was very similar in most of the scenarios with a slightly better performance for the stratified model for small cluster size at short time horizons. Third, the frailty Cox model tended to have better calibration than the stratified Cox for low number of clusters with small size and low homogeneity within clusters at short time horizons. Calibration tended to be similar in both models as size and number of clusters increased especially with longer time horizons. Across all scenarios, the stratified and frailty Cox had generally similar performances.

To the best of our knowledge, this is the first study that investigates the impact of number, size and homogeneity within clusters on the prediction performance of Cox proportional hazards models for censored time-to-event clustered data using simulations. The Cox proportional hazards model is the most popular regression model for time-to-event outcomes. The Cox proportional hazards model may manage clusters non parametrically using stratification or parametrically using the frailty (i.e., random effect) term. Thus, the stratified approach makes less assumptions compared to the frailty approach, although the former approach can be less efficient and less useful when the size of the cluster is small^33^. To our knowledge, other works primarily focused on estimating the effect of the intervention in multicentre (randomized) clinical trials^34–36^. Glidden and Vittinghoff investigated how to incorporate centres effects into the analysis of multicentre clinical trials for censored time-to-event outcomes^34^. They found substantial advantages of the gamma frailty Cox model compared to the fixed and stratified Cox model. Munda and Legrand extended the findings of Glidden and Vittinghoff also when the frailty distribution is misspecified^35^. de Jong et al. focused on the one stage approach (i.e., all subjects from different studies are analysed in a single step) and two stage approach (i.e., subjects are analysed separately in each study and then results are aggregated) for IPD-MA of intervention studies with time-to-event outcomes^36–38^. The authors suggested similar conclusions with previous works. While these prior studies focused on estimating intervention effects in RCTs, less is known about the impact of clustered data on prediction performances of risk prediction models for survival outcomes. In the one stage approach of IPD-MA for intervention and prognostic studies with time-to-event outcomes, Riley and colleagues stated “It is fundamental to account for clustering using either stratified or frailty models […]. Frailty models are perhaps best when the number of participants and outcome events per trial are small, given the estimation issues may arise with separate baseline hazards per trial”^32,39^. With this simulation study, we aimed to give further insights for applied researchers. We essentially confirmed the statement of Riley and colleagues and the literature’s findings about the multicentre interventional studies^32,34–36^. Our simulation study suggested that prediction performance was generally similar across most of the investigated scenarios. For small cluster size with higher number of clusters and moderate to high homogeneity within clusters, the frailty and stratified model tended to results in similar prediction performances with slightly better discrimination and overall measures for the stratified model. The frailty model tended to result in better calibration for small cluster size especially for mild within-cluster homogeneity and at short time horizons. However, the practical utility of the stratified Cox proportional hazards model is limited since the baseline hazard in every cluster and the corresponding estimated risk predictions cannot be always estimated for a considerable heterogeneous number of clusters with small size and in particular at longer time horizons.

Our study had several limitations. First, our study relied on a limited number of scenarios for the Monte Carlo simulations. This was mainly due to computationally intensive simulations. On the other hand, we provided results from 80 scenarios for three time horizons for a total of 240 scenarios. Secondly, we generated event times using the Cox-Weibull framework^11^. Different models could have been investigated to generate outcomes. Thirdly, our work focused exclusively on predictions in the same clusters used to develop the risk prediction model. Therefore, our study did not consider marginal prediction performance assessment of the marginal gamma frailty Cox model. It is formally not possible to provide marginal predictions from the stratified Cox model, unless an appropriate baseline hazard could then be selected from existing studies in the meta-analysis using the incidence in the new study population^31^. In the context of IPD-MA, the prediction performance assessment is generally evaluated using the internal-external cross validation^31,40^.

An important limitation of the stratified Cox model was the impossibility to estimate the predictions for all clusters. The stratified Cox models estimate a baseline hazard for every stratum (i.e., cluster). Cluster-specific (i.e., stratum-specific) baseline hazard could not be estimated for clusters in which time horizons were greater than the maximum observed event time of the corresponding clusters in the derivation sample. Thus, the corresponding predictions from the stratified Cox model could not be estimated. Therefore, this substantially limits the utility of the stratified Cox compared to the frailty Cox model. In our simulations, the number of usable comparisons between the two models decreased substantially for longer time horizons and higher homogeneity within clusters (especially among scenarios with higher number of clusters with small size). The scenario with the least relative utility of the comparisons between the two models was observed for a homogeneity within clusters of 0.10 with 8 clusters and with an average number of 25 patients per cluster where only 14.1% of comparisons between the two models were possible (details available in the supplementary). In the frailty Cox model, a shared baseline hazard is estimated across clusters which enables to provide predictions over a considerably larger range of reasonable potential prediction time horizons.

In conclusion, the stratified and frailty Cox proportional hazards models generally provide similar predictions performances in most of the simulated scenarios with different number and sizes of clusters. When the cluster size is small, the stratified Cox proportional hazard model provides slightly better discrimination but worse calibration compared to the shared gamma frailty Cox proportional hazards model especially at short time horizons and when homogeneity within clusters is mild. The practical applicability of the stratified Cox proportional hazards model to estimate predictions is limited especially for high heterogeneity between clusters and when clusters are small, and more likely at longer time prediction horizons. The shared gamma frailty Cox proportional hazards model can provide predictions in case of small and heterogeneous clusters. It also enables marginal predictions for individuals from new or previously unobserved clusters.

## Data Availability

All data produced are available online at https://github.com/danielegiardiello/ClusterSurvPred

https://github.com/danielegiardiello/ClusterSurvPred/blob/main/Data/insem.rds

## Acknowledgments

We thank Stefano Bottelli for his suggestions on this manuscript.

## Data availability Statement

All the R code and datasets used to run the case studies is freely available online at https://github.com/danielegiardiello/ClusterSurvPred. The first case study dataset was collected by the Center for International Blood and Marrow Transplant Research (CIBMTR) which is supported primarily by the Public Health Service U24CA076518 from the National Cancer Institute; the National Heart, Lung, and Blood Institute; the National Institute of Allergy and Infectious Diseases; 75R60222C00011 from the Health Resources and Services Administration; N00014-23-1-2057 and N00014-24-1-2507 from the Office of Naval Research; NMDP; and the Medical College of Wisconsin.

---

**Figure S1.**
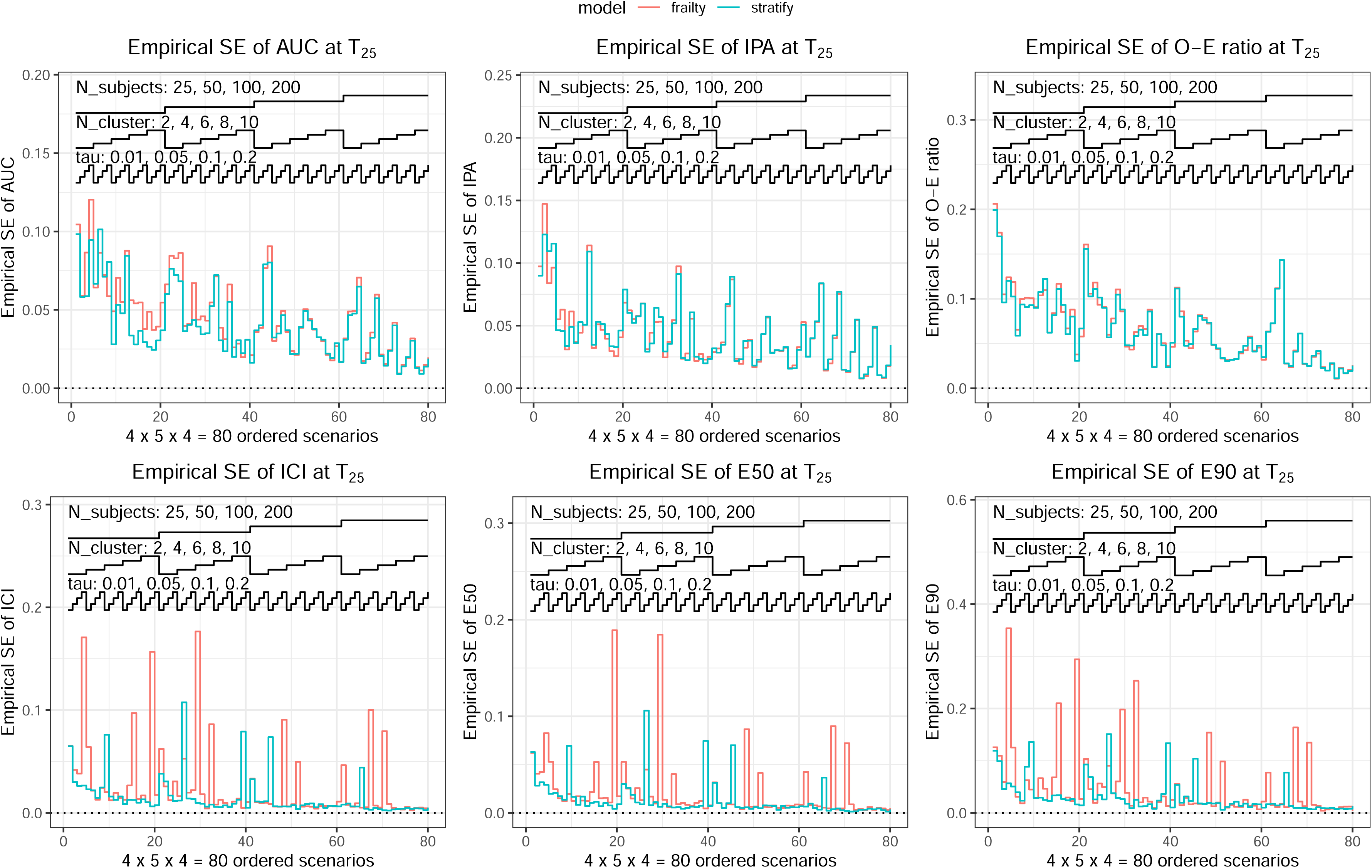
SE of the prediction performances at T_25_.

**Figure S2.**
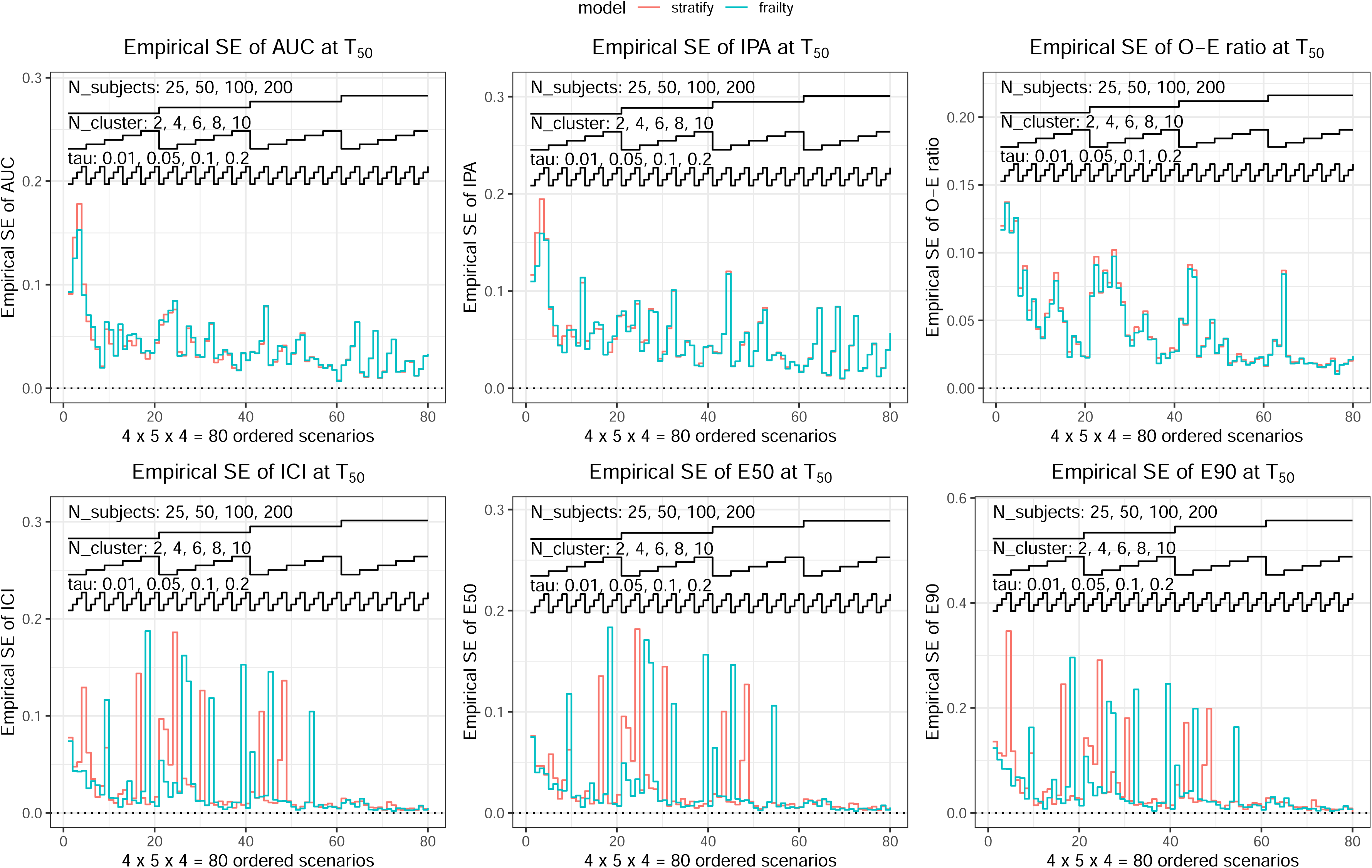
SE of the prediction performances at T_50_.

**Figure S3.**
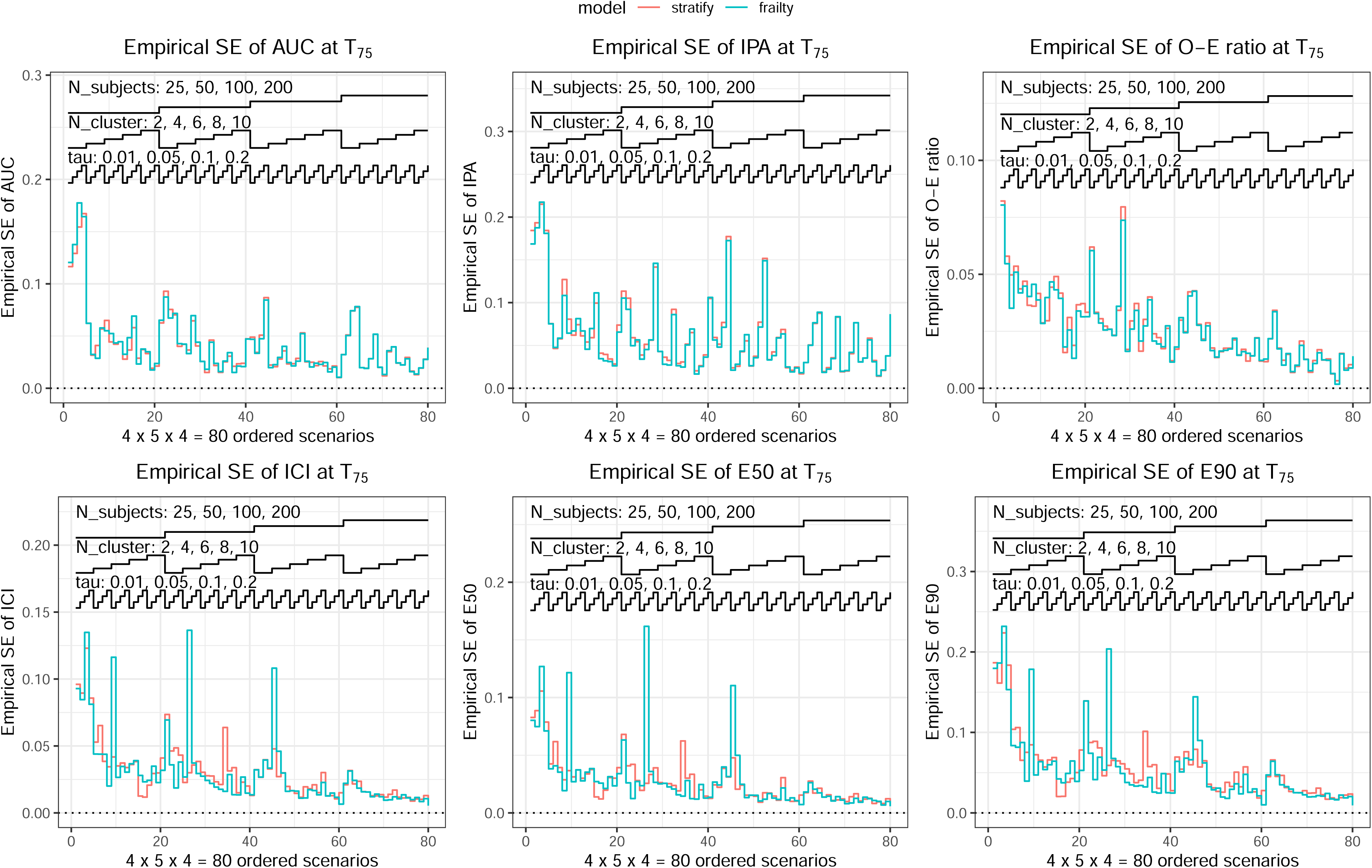
SE of the prediction performances at T_75_.

## References

1. Majhail NS, Mau LW, Chitphakdithai P, et al. National Survey of Hematopoietic Cell Transplantation Center Personnel, Infrastructure, and Models of Care Delivery. Biol Blood Marrow Transplant. 2015;21(7):1308–1314.

2. Frassoni F, Labopin M, Powles R, et al. Effect of centre on outcome of bone-marrow transplantation for acute myeloid leukaemia. Acute Leukaemia Working Party of the European Group for Blood and Marrow Transplantation. Lancet. 2000;355(9213):1393–1398.

3. Loberiza FR, Zhang MJ, Lee SJ, et al. Association of transplant center and physician factors on mortality after hematopoietic stem cell transplantation in the United States. Blood. 2005;105(7):2979–2987.

4. Majhail NS, Mau LW, Chitphakdithai P, et al. Transplant Center Characteristics And Survival After Allogeneic Hematopoietic Cell Transplantation In Adults. Bone Marrow Transplant. 2020;55(5):906–917.

5. Cox, DR. Regression Models and Life Tables (With Discussion). *Journal of the Royal Statistical Society*, Series B (Statistical Methodology). 1972;34:187–202.

6. Steyerberg EW, Vickers AJ, Cook NR, et al. Assessing the performance of prediction models: a framework for traditional and novel measures. Epidemiology. 2010;21(1):128–138.

7. Wynants L, Vergouwe Y, Van Huffel S, Timmerman D, Van Calster B. Does ignoring clustering in multicenter data influence the performance of prediction models? A simulation study. Stat Methods Med Res. 2018;27(6):1723–1736.

8. Falconieri N, Van Calster B, Timmerman D, Wynants L. Developing risk models for multicenter data using standard logistic regression produced suboptimal predictions: A simulation study. Biom J. 2020;62(4):932–944.

9. Duchateau, L, Janssen, P. The Frailty Model. First. Springer New York, NY; 2008.

10. Wienke A. Frailty Model in Survival Analysis. First. Chapman & Hall/CRC Press; 2011.

11. Bender R, Augustin T, Blettner M. Generating survival times to simulate Cox proportional hazards models. Statistics in Medicine. 2005;24.

12. Hougaard, P. Analysis of Multivariate Survival Data. 1st ed. Springer New York, NY; 2000.

13. Austin PC. The iterative bisection procedure: a useful tool for determining parameter values in data-generating processes in Monte Carlo simulations. BMC Medical Research Methodology. 2023;23(1):45.

14. Gerds TA, Kattan MW. Medical Risk Prediction Models: With Ties to Machine Learning. 1st ed. Chapman & Hall/CRC Press; 2022.

15. McLernon DJ, Giardiello D, Van Calster B, et al. Assessing Performance and Clinical Usefulness in Prediction Models With Survival Outcomes: Practical Guidance for Cox Proportional Hazards Models. Ann Intern Med. 2023;176(1):105–114.

16. Blanche P, Kattan MW, Gerds TA. The c-index is not proper for the evaluation of t-year predicted risks. Biostatistics. 2019;20(2):347–357.

17. Kattan MW, Gerds TA. The index of prediction accuracy: an intuitive measure useful for evaluating risk prediction models. Diagnostic and Prognostic Research. 2018;2(1):7.

18. Austin, PC, Harrell, FE, van Klaveren D. Graphical calibration curves and the integrated calibration index (ICI) for survival models. Stat Med. 2020;39(21):2714–2742.

19. Kooperberg C, Stone CJ, Truong YK. Hazard Regression. Journal of the American Statistical Association. Published online 1995.

20. R Core Team. R: A language and environment for statistical computing.

21. Therneau, MT, Grambsch, PM. Frailty Models (chapter 9). In: Modeling Survival Data - Extending the Cox Model. Statistics for Biology and Health. Springer New York, NY.

22. Therneau TM, Grambsch PM, Pankratz VS. Penalized Survival Models and Frailty. Journal of Computational and Graphical Statistics. 2003;12(1):156–175.

23. Rücker G, Schwarzer G. Presenting simulation results in a nested loop plot. BMC Medical Research Methodology. 2014;14(1):129.

24. Horowitz M. The role of registries in facilitating clinical research in BMT: examples from the Center for International Blood and Marrow Transplant Research. Bone Marrow Transplant. 2008;42 Suppl 1:S1–S2.

25. Nazha A, Hu ZH, Wang T, et al. A Personalized Prediction Model for Outcomes after Allogeneic Hematopoietic Cell Transplant in Patients with Myelodysplastic Syndromes. Biol Blood Marrow Transplant. 2020;26(11):2139–2146.

26. Greenberg P, Cox C, LeBeau MM, et al. International scoring system for evaluating prognosis in myelodysplastic syndromes. Blood. 1997;89(6):2079–2088.

27. Greenberg PL, Tuechler H, Schanz J, et al. Revised international prognostic scoring system for myelodysplastic syndromes. Blood. 2012;120(12):2454–2465.

28. Shaffer BC, Ahn KW, Hu ZH, et al. Scoring System Prognostic of Outcome in Patients Undergoing Allogeneic Hematopoietic Cell Transplantation for Myelodysplastic Syndrome. J Clin Oncol. 2016;34(16):1864–1871.

29. Sylvester RJ, van der Meijden APM, Oosterlinck W, et al. Predicting recurrence and progression in individual patients with stage Ta T1 bladder cancer using EORTC risk tables: a combined analysis of 2596 patients from seven EORTC trials. Eur Urol. 2006;49(3):466–465; discussion 475-477.

30. Ahmed I, Debray TP, Moons KG, Riley RD. Developing and validating risk prediction models in an individual participant data meta-analysis. BMC Medical Research Methodology. 2014;14(1):3.

31. Debray TPA, Moons KGM, Ahmed I, Koffijberg H, Riley RD. A framework for developing, implementing, and evaluating clinical prediction models in an individual participant data meta-analysis. Stat Med. 2013;32(18):3158–3180.

32. Riley, RD, Tierney, J, Stewart, LA (Eds). Individual Participant Data Meta_-_Analysis: A Handbook for Healthcare Research. John Wiley & Sons Ltd; 2021.

33. O’Quigley, J. Proportional Hazards Regression. Springer New York, NY; 2008.

34. Glidden DV, Vittinghoff E. Modelling clustered survival data from multicentre clinical trials. Stat Med. 2004;23(3):369–388.

35. Munda M, Legrand C. Adjusting for centre heterogeneity in multicentre clinical trials with a timeltolevent outcome. Pharmaceutical Statistics. 2014;13(2):145–152.

36. de Jong VMT, Moons KGM, Riley RD, et al. Individual participant data meta-analysis of intervention studies with time-to-event outcomes: A review of the methodology and an applied example. Research Synthesis Methods. 2020;11(2):148–168.

37. Burke DL, Ensor J, Riley RD. Meta-analysis using individual participant data: one-stage and two-stage approaches, and why they may differ. Stat Med. 2017;36(5):855–875.

38. Riley RD, Ensor J, Hattle M, Papadimitropoulou K, Morris TP. Two-stage or not two-stage? That is the question for IPD meta-analysis projects. Research Synthesis Methods. 2023;14(6):903–910.

39. Riley, RD, Debray, TPA. The One-stage Approach to IPD Meta-analysis (chapter 6, section 6.3). In: Individual Participant Data Meta-Analysis: A Handbook for Healthcare Research. John Wiley & Sons Ltd; 2021.

40. Steyerberg EW, Harrell FE. Prediction models need appropriate internal, internal-external, and external validation. J Clin Epidemiol. 2016;69:245–247.

