## Supplementary material for "The impact of the number and the size of clusters on prediction performance of the stratified and the conditional shared gamma frailty Cox proportional hazards models"

Supplementary Table 1. Percentage (%) of usable comparisons across the 37 of 80 scenarios in which the prediction performance of the stratified Cox proportional hazards model could not be assessed in every cluster across all 1,000 iterations per scenario.

| **N_subjects_** | **N_clusters_** | $\boldsymbol{\tau}$ | **Times** | **%** |
| --- | --- | --- | --- | --- |
| 25 | 8 | 0.10 | *T_75_* | 14.1 |
| 100 | 8 | 0.20 | *T_75_* | 30.6 |
| 25 | 8 | 0.20 | *T_75_* | 35.3 |
| 25 | 10 | 0.20 | *T_75_* | 46.6 |
| 200 | 8 | 0.20 | *T_75_* | 49.2 |
| 25 | 10 | 0.01 | *T_75_* | 56.2 |
| 25 | 2 | 0.10 | *T_75_* | 60.4 |
| 100 | 10 | 0.20 | *T_75_* | 63.2 |
| 25 | 4 | 0.20 | *T_75_* | 63.4 |
| 25 | 10 | 0.05 | *T_75_* | 65.3 |
| 25 | 6 | 0.20 | *T_75_* | 67.3 |
| 50 | 8 | 0.20 | *T_75_* | 67.4 |
| 50 | 10 | 0.20 | *T_75_* | 67.5 |
| 25 | 8 | 0.05 | *T_75_* | 69.4 |
| 25 | 4 | 0.10 | *T_75_* | 75.0 |
| 25 | 4 | 0.01 | *T_75_* | 76.3 |
| 50 | 6 | 0.10 | *T_75_* | 76.5 |
| 25 | 4 | 0.05 | *T_75_* | 79.8 |
| 25 | 8 | 0.01 | *T_75_* | 79.9 |
| 50 | 8 | 0.10 | *T_75_* | 81.3 |
| 100 | 8 | 0.05 | *T_75_* | 81.5 |
| 25 | 2 | 0.05 | *T_75_* | 82.1 |
| 50 | 10 | 0.10 | *T_75_* | 83.4 |
| 25 | 2 | 0.20 | *T_75_* | 83.4 |
| 25 | 10 | 0.20 | *T_50_* | 83.4 |
| 100 | 8 | 0.10 | *T_75_* | 85.9 |
| 200 | 10 | 0.20 | *T_75_* | 85.9 |
| 25 | 10 | 0.10 | *T_75_* | 86.0 |
| 50 | 4 | 0.20 | *T_75_* | 86.0 |
| 50 | 6 | 0.20 | *T_75_* | 86.0 |
| 25 | 6 | 0.01 | *T_75_* | 88.4 |
| 25 | 6 | 0.05 | *T_75_* | 88.8 |
| 25 | 2 | 0.01 | *T_75_* | 91.3 |
| 50 | 6 | 0.05 | *T_75_* | 92.7 |
| 25 | 6 | 0.10 | *T_75_* | 92.8 |
| 50 | 8 | 0.01 | *T_75_* | 92.9 |
| 50 | 6 | 0.10 | *T_50_* | 93.1 |

Supplementary Table 2: descriptive statistics of the number of the usable iterations (out of 1 000 iterations per scenario) by prediction time horizons

| Time horizon | Minimum | 25^th^ percentile | 50^th^ percentile | Mean | 75^th^ percentile | Maximum |
| --- | --- | --- | --- | --- | --- | --- |
| *T_25_* | 1000 | 1000 | 1000 | 1000 | 1000 | 1000 |
| *T_50_* | 834 | 1000 | 1000 | 997 | 1000 | 1000 |
| *T_75_* | 141 | 814 | 1000 | 879 | 1000 | 1000 |

| Supplementary Table 3: comparison of stratified versus frailty Cox proportional hazards models on prediction performance metrics across all comparable simulation scenarios and by time horizons | | | | | | | |
| --- | --- | --- | --- | --- | --- | --- | --- |
| **Performance  metric** | **Time**  **horizon** | **Comparison** | **Minimum** | **25^th^  percentile** | **50^th^  percentile** | **75^th^  percentile** | **Maximum** |
| AUC | *T_25_* | Stratify vs frailty | -0.009 | 0.002 | 0.005 | 0.015 | 0.033 |
| AUC | *T_50_* | Stratify vs frailty | -0.018 | 0.000 | 0.001 | 0.003 | 0.014 |
| AUC | *T_75_* | Stratify vs frailty | -0.022 | -0.001 | 0.000 | 0.002 | 0.017 |
| IPA | *T_25_* | Stratify vs frailty | -0.007 | 0.000 | 0.003 | 0.007 | 0.024 |
| IPA | *T_50_* | Stratify vs frailty | -0.015 | 0.001 | 0.000 | 0.003 | 0.013 |
| IPA | *T_75_* | Stratify vs frailty | -0.027 | -0.003 | -0.001 | 0.001 | 0.032 |
| O-E ratio | *T_25_* | Stratify vs frailty | -0.023 | -0.003 | 0.003 | 0.010 | 0.031 |
| O-E ratio | *T_50_* | Stratify vs frailty | -0.023 | -0.003 | 0.002 | 0.013 | 0.027 |
| O-E ratio | *T_75_* | Stratify vs frailty | -0.032 | -0.004 | 0.003 | 0.009 | 0.032 |
| ICI | *T_25_* | Stratify vs frailty | -0.032 | 0.002 | 0.005 | 0.012 | 0.093 |
| ICI | *T_50_* | Stratify vs frailty | -0.084 | -0.001 | 0.002 | 0.006 | 0.100 |
| ICI | *T_75_* | Stratify vs frailty | -0.106 | -0.001 | 0.001 | 0.005 | 0.048 |
| E50 | *T_25_* | Stratify vs frailty | -0.035 | 0.002 | 0.0053 | 0.011 | 0.101 |
| E50 | *T_50_* | Stratify vs frailty | -0.084 | -0.002 | -0.0013 | 0.006 | 0.093 |
| E50 | *T_75_* | Stratify vs frailty | -0.136 | -0.001 | -0.0013 | 0.004 | 0.048 |
| E90 | *T_25_* | Stratify vs frailty | -0.056 | 0.001 | 0.008 | 0.020 | 0.183 |
| E90 | *T_50_* | Stratify vs frailty | -0.128 | -0.003 | -0.003 | 0.011 | 0.150 |
| E90 | *T_75_* | Stratify vs frailty | -0.145 | -0.002 | -0.003 | 0.011 | 0.069 |
| AUC: area Under the receiver operating characteristic curve Curve; O-E: observed-to-expected; ICI: Integrated Calibration Index | | | | | | | |
